# Cross-classification between self-rated health and health status: longitudinal analyses of all-cause mortality and leading causes of death in the UK

**DOI:** 10.1101/2021.04.23.21255982

**Authors:** Julian Mutz, Cathryn M. Lewis

## Abstract

**Background:** Risk stratification is an important public health priority that is central to clinical decision making and resource allocation. The aim of the present study was to examine how different combinations of self-rated and objective health status predict (i) all-cause mortality and (ii) cause-specific mortality from leading causes of death in the UK.

**Methods:** The UK Biobank study recruited >500,000 participants, aged 37-73, between 2006–2010. The health cross-classification examined incorporated self-rated health (poor, fair, good or excellent) and health status derived from medical history and current disease status, including 81 cancer and 443 non-cancer illnesses. We examined all-cause mortality and six specific causes of death: ischaemic heart disease, cerebrovascular disease, influenza and pneumonia, dementia and Alzheimer’s disease, chronic lower respiratory disease and malignant neoplasm.

**Results:** Analyses included **>**370,000 middle-aged and older adults with a median follow-up of 11.75 (IQR = 1.4) years, yielding 4,320,270 person years of follow-up. Compared to excellent self-rated health and favourable health status, all other levels of the health cross-classification were associated with a greater risk of mortality, with hazard ratios ranging from 1.22 (95% CI 1.15-1.29, *p*_Bonf._ < 0.001) for good self-rated health and favourable health status to 7.14 (95% CI 6.70-7.60, *p*_Bonf._ < 0.001) for poor self-rated health and unfavourable health status.

**Conclusions:** Our findings highlight that self-rated health captures additional health-related information and should be more widely assessed across settings. The cross-classification between health status and self-rated health represents a straightforward metric for risk stratification, with applications to population health, clinical decision making and resource allocation.

## Introduction

Self-rated health is used extensively in epidemiological and public health research, and a plethora of studies have found it to be predictive of morbidity and mortality^1^. It constitutes a single item measure of subjective health status that likely encompasses biological, psychological and social dimensions.

Despite its simplicity, self-rated health is correlated with objective assessments of health^2^. For example, research in more than 16,000 Chinese residents aged 18-80 found a higher prevalence of diseases and abnormalities in laboratory tests in individuals with poor self-rated health^3^. A study of 1,322 community-dwelling elderly aged 60 or older who participated in the Bambui Cohort Study of Aging in Brazil examined how well self-rated health predicted 10-year mortality, compared to a comprehensive health score derived from objective clinical measures^4^. Individuals with poor self-rated health had a two-fold increased risk of death during the follow-up period, compared to individuals who rated their health as good or excellent. Self-rated health was comparable to the comprehensive health score in predicting mortality and remained predictive of mortality after adjustment for the health score, suggesting that it captures additional health-related information.

Electronic health records and other patient data that are available to health care providers could be used to derive a measure of overall health status. Self-rated health can be assessed using a single-item question and is for instance included in census questionnaires^5^ but not, to our knowledge, routinely collected during primary care registrations or visits. The cross-classification between self-rated health and objective health status based on medical history or clinical measures could represent a readily available metric for risk stratification. The identification of at-risk populations is an important public health priority that is central to clinical decision making and resource allocation.

The aim of the present study was to examine the relative contributions of self-rated health and health status based on medical history and current disease status to predict mortality in middle-aged and older adults in the UK Biobank. We created a matrix of the cross-classification between self-rated health and health status and examined how different combinations of self-rated and objective health status predict (i) all-cause mortality and (ii) cause-specific mortality from leading causes of death in the UK^6^ during a follow-up period of approximately 12 years.

We hypothesised that individuals with concordant self-rated health and health status would demonstrate the longest (favourable health status and good or excellent self-rated health) and shortest (unfavourable health status and poor or fair self-rated health) survival times, respectively. Individuals with discordant self-rated health and health status were predicted to demonstrate intermediate survival times, although no predictions were made about the precise order of survival times (Supplement 1).

The aim of comparing these survival times was to explore whether the discordant categories would provide additional information about the relative importance of self-rated health and health status with respect to future health outcomes.

## Methods

### Study population

The UK Biobank is a prospective study of > 500,000 UK residents aged 37–73 at baseline, recruited between 2006–2010. Details of the study rationale and design have been reported elsewhere^7^. Briefly, individuals registered with the UK National Health Service (NHS) and living within a 25-mile (∼40 km) radius of one of 22 assessment centres were invited to participate (9,238,453 postal invitations sent). At the baseline assessment, participants completed electronic questionnaires and nurse-led interviews to provide data on sociodemographic characteristics, health behaviours and their medical history. Linked hospital inpatient records are available for most participants and these data have been linked to death registries.

### Exposures

Data on 81 cancer and 443 non-cancer illnesses (past and current) were ascertained through touchscreen self-report questionnaire and confirmed during a verbal interview by a trained nurse. In order to provide a single health indicator (“health status”) based on a previously defined algorithm, we used a classification developed by the Reinsurance Group of America (RGA) in which an experienced underwriter classified each illness according to whether it was “likely acceptable for standard life insurance”^8^. Participants were thus classified as having favourable or unfavourable health status based on their reported cancer and non-cancer illnesses. Details of this classification have been reported previously^8,9^.

Participants’ self-rated health was assessed using the question “In general how would you rate your overall health?”. Response options included “Poor”, “Fair”, “Good” and “Excellent”.

We derived a measure of the cross-classification between health status and self-rated health. Individuals with missing data or who responded “prefer not to answer” or “do not know” to the self-rated health question were excluded.

### Ascertainment of mortality

Our primary outcome was all-cause mortality, i.e., death from any cause. All-cause mortality represents a standard index used in clinical decision making that is easily assessed and concrete^10^. The date of death was obtained through linkage with national death registries from NHS Digital for participants in England and Wales and from the NHS Central Register for participants in Scotland.

The censoring date for mortality was 30 November 2020. The most recent death was recorded for 18 December 2020, although data were not complete for December 2020.

We also examined cause-specific mortality for leading causes of death in the UK^6^. The primary cause of death was recorded based on the International Classification of Diseases (10th revision). The following outcomes were examined: ischaemic heart diseases (I20-I25), cerebrovascular diseases (I60-I69), influenza and pneumonia (J09-J18), dementia and Alzheimer’s disease (F01, F03 and G30), chronic lower respiratory diseases (J40-J47) and malignant neoplasms (C00-C97). For each cause-specific death, individuals who died of other causes were censored at their age at death.

### Covariates

Potential confounders of the association between self-rated health or health status and all-cause or cause-specific mortality were identified from the baseline assessment data: sociodemographic factors (sex, ethnicity [6 levels], highest educational/professional qualification [4 levels]^11^ and annual household income [5 levels]), lifestyle (smoking [3 levels], alcohol intake frequency [6 levels], physical activity [number of days per week spent walking, engaging in moderate-intensity physical activity or engaging in vigorous-intensity physical activity for ≥10 minutes continuously]). All covariates were assumed to be fixed.

### Exclusion criteria

Participants for whom their genetic sex, inferred from the genotype information on the Y and X chromosomes, and self-reported sex did not match were excluded. Individuals with missing data or who responded “do not know” or “prefer not to answer” to any of the assessed covariates were also excluded from analyses.

### Statistical analyses

All analyses were pre-specified prior to inspection of the data (preregistration: osf.io/qvm39) and algorithms were tested on simulated data. Statistical analyses were conducted using R (version 3.6.0).

Characteristics of the full and analytical sample were summarised using means and standard deviations or counts and percentages. The total number of self-reported diseases and the frequencies of illnesses by disease group were summarised for each level of the cross-classification between health status and self-rated health.

We present the number of individuals who died during follow-up of any cause (all-cause mortality) and of specific causes (cause-specific mortality). The minimum number of observed deaths was set a priori at 20 for each level of our primary exposure. This criterion was based on a previous UK Biobank study of 5-year mortality^12^. Finally, we calculated person-years of follow-up and the median duration of follow-up of censored individuals.

Unadjusted survival probabilities by health status, self-rated health and health cross-classification were estimated non-parametrically from observed survival times using the Kaplan-Meier (KM) method^13^. We present KM survival curves and *p*-values from log-rank tests. Hazard ratios (HRs) and 95% confidence intervals were estimated using Cox proportional hazards models^14^ to examine associations between the health cross-classification and mortality adjusted for potential confounders. Age (in years) was used as the underlying time axis, conditional on living to age 40. We fitted a sequence of three models: Model 1 – unadjusted; Model 2 – adjusted for sociodemographic characteristics; Model 3 – additionally adjusted for lifestyle factors. We present plots of the estimated survival probability and cumulative hazards. Continuous covariates were fixed at their mean value while categorical covariates were fixed at the largest group. When presenting results in tables and figures, the levels of the health cross-classification are shown in ascending order by their HR for all-cause mortality from the results of Model 1. Scaled Schoenfeld residuals and Log(-log(survival)) curves as a function of time were examined to assess the assumption of proportional hazards.

Martingale residuals were visually inspected to assess the assumption of log-linearity for continuous covariates.

Adjusted *p*-values were calculated using the p.adjust() command in R to account for multiple testing across models, separately for each outcome. *P*-values were corrected for 21 tests (three models × seven estimated parameters). Two methods were used: (1) Bonferroni and (2) Benjamini & Hochberg^15^, all two-tailed, with *α* = .05 and false discovery rate of 5%, respectively.

### Sensitivity analysis

In a sensitivity analysis we examined a simplified health cross-classification in which fair and poor self-rated health and good and excellent were merged prior to analysis (**Supplement 1**). Self-rated health levels have been combined previously, although with 5 initial response options^16^.

## Results

### Sample characteristics

Of the 502,521 UK Biobank participants, 487,195 (96.95%) had complete data on health cross-classification. We retained an analytical sample of 373,761 participants after removing individuals with missing data on covariates (*n* = 118,190), who did not meet our inclusion criteria (*n* = 372) or whose recorded date of death was before or on the same day as the baseline assessment (*n* = 495) (Figure 1).

**Figure 1.**
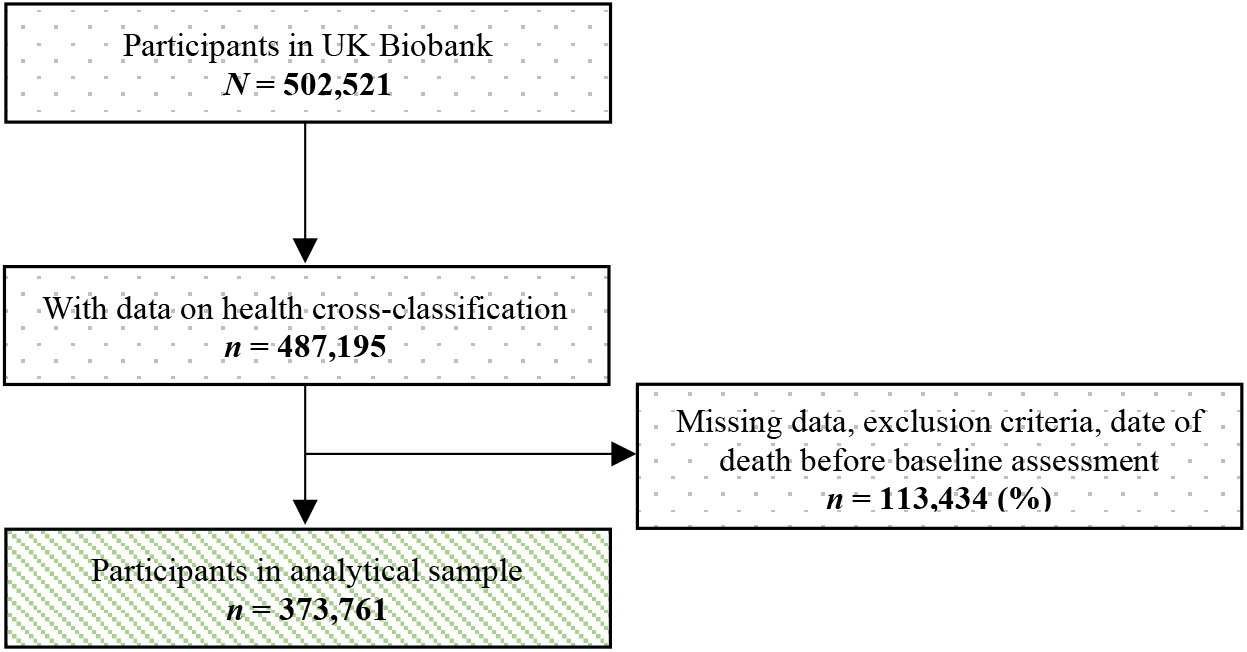
Flowchart of study sample.

Descriptive statistics of the full and analytical samples are presented in Supplement Table 1. The mean age at baseline assessment of participants included in our analytical sample was 56.02 (SD = 8.08) years and 51.8% of these participants were female. We classified 117,212 (31.36%) participants as having an unfavourable health status and 256,549 (68.64%) participants as having a favourable health status. Most participants (*n* = 219,628, 58.76%) had good self-rated health and 14,185 (3.80%), 73,138 (19.57%) and 66,810 (17.88%) participants had poor, fair and excellent self-rated health, respectively (Table 1).

**Table 1.** Cross-classification of health status and self-rated health

| Health status | Self-rated health |  |  |  |
| --- | --- | --- | --- | --- |
| | Poor<br>$N = 14185$ | Fair<br>$n = 73138$ | Good<br>$n = 219628$ | Excellent<br>$n = 66810$ |
| Favourable<br>$n = 256549$ | 4004<br>(1.07%) | 37863<br>(10.13%) | 159173<br>(42.59%) | 55509<br>(14.85%) |
| Unfavourable<br>$n = 117212$ | 10181<br>(2.72%) | 35275<br>(9.44%) | 60455<br>(16.17%) | 11301<br>(3.02%) |

Note: Percentages in cells are based on complete analytical sample of $n = 373,761$ .

The average number of illnesses by health cross-classification ranged from 0.73 (SD = 0.99) in individuals with excellent self-rated health and favourable health status to 4.93 (SD = 2.70) in individuals with poor self-rated health and unfavourable health status (Table 2). The distributions of the number of illnesses by health cross-classification are presented in Figure 2 and show that there were substantial differences between groups. The proportions of individuals with at least one cancer or non-cancer illness within several broader illness groups are presented by health cross classification in **Supplementary Figure 1** and **Supplementary Figure 2**.

**Table 2.**
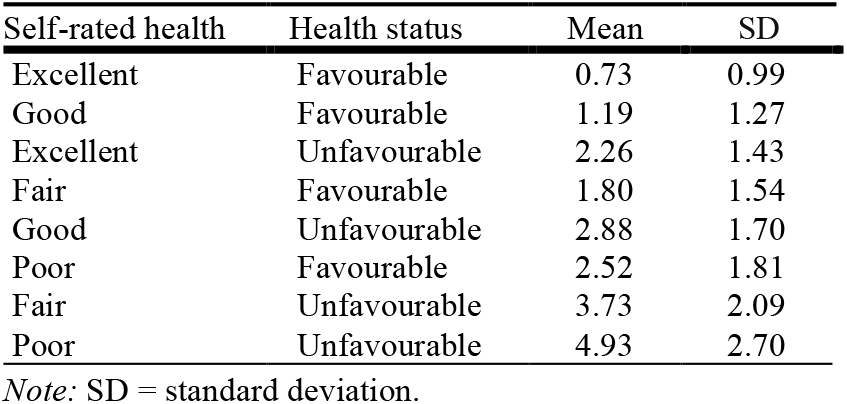
Number of illnesses by health cross-classification

**Figure 2.**
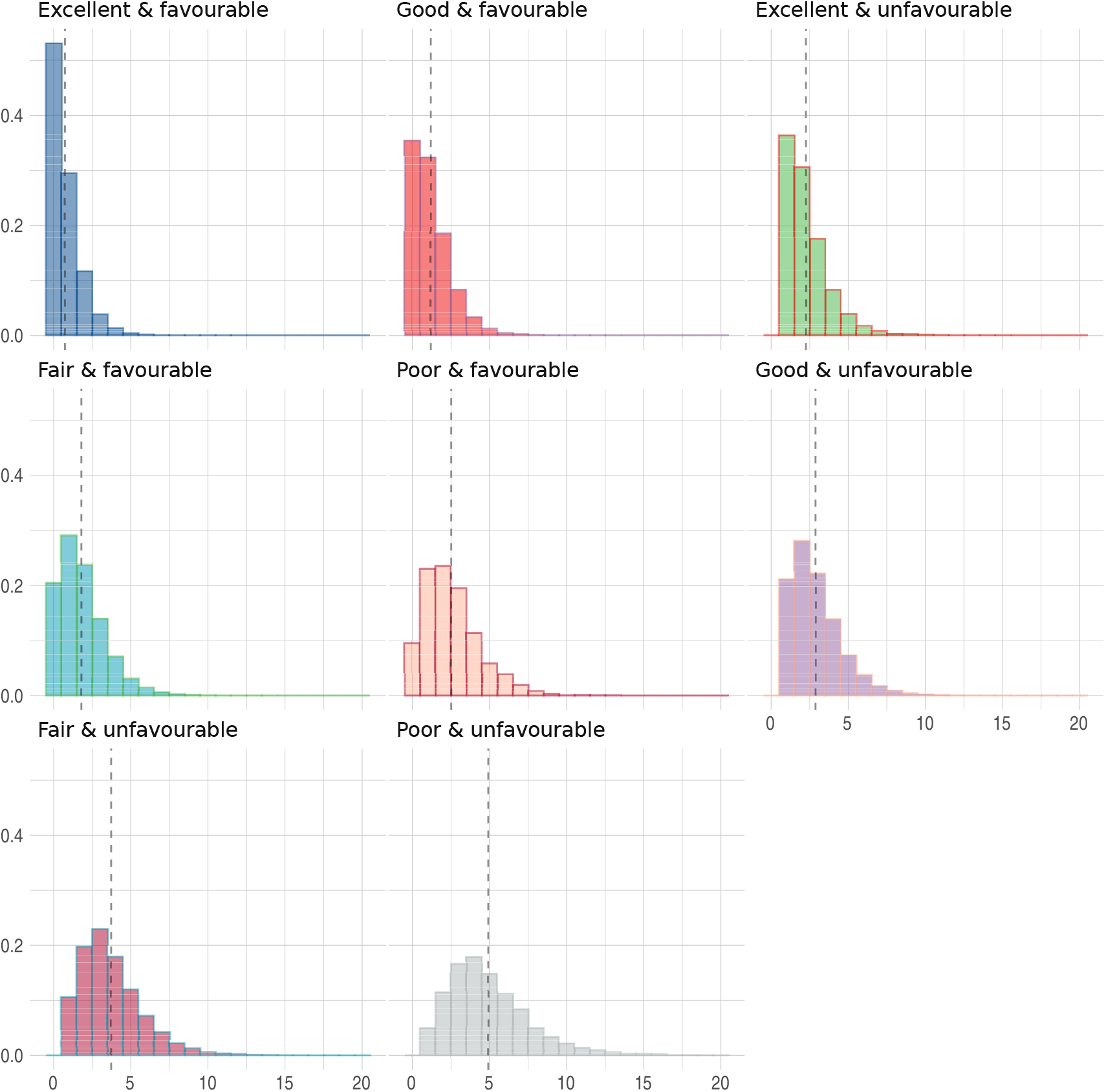
Number of illnesses by health cross-classification. 9 observations were above graph maximum of 20 illnesses (5 poor & unfavourable, 3 fair & unfavourable and 1 excellent & unfavourable).

The median follow-up was 11.75 (IQR = 1.4) years, yielding 4,320,270 person years of follow-up. The median follow-up of censored individuals was 11.81 (IQR = 1.35) years and the potential median survival time, calculated using the reverse Kaplan-Meier method, was 11.819 days (95% CI = 11.814-11.822). We observed 21,980 (5.88%) deaths from any cause. Deaths observed for specific causes were 2,432 (0.65%) for ischaemic heart disease, 908 (0.24%) for cerebrovascular disease, 409 (0.11%) for influenza and pneumonia, 701 (0.19%) for dementia and Alzheimer’s disease, 699 (0.19%) for chronic lower respiratory disease and 11,171 (2.99%) for malignant neoplasms.

### All-cause mortality

Kaplan-Meier survival probabilities for all-cause mortality are presented in Figure 3. Favourable health status and better self-rated health were associated with longer survival times (log-rank test *p*-values < 0.001). Different levels of the cross-classification between health status and self-rated health were associated with varying survival times (log-rank test *p* < 0.001). We found no evidence that survival probabilities differed by year of attending the baseline assessment (*p* = 0.15).

**Figure 3.**
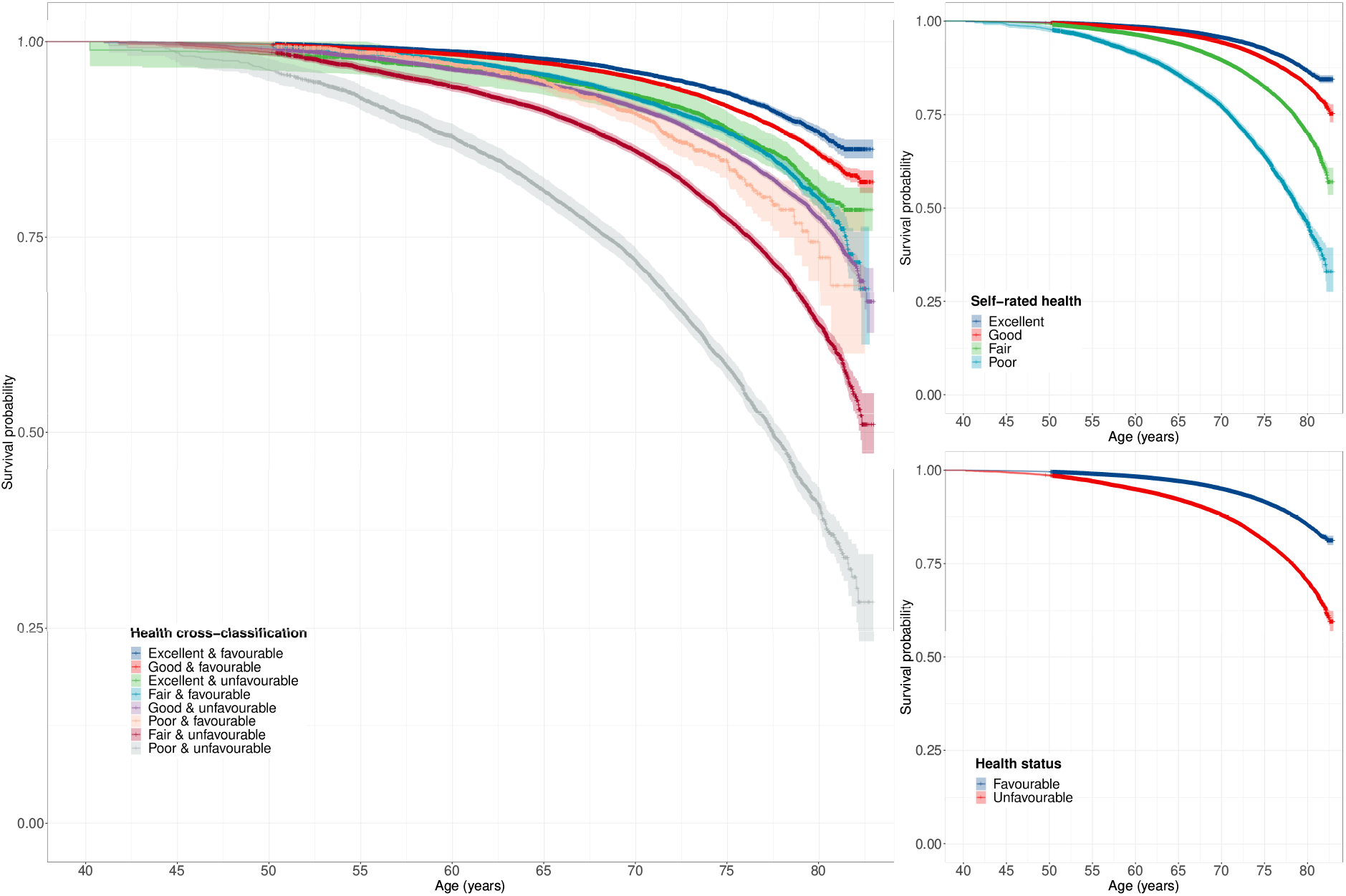
Kaplan-Meier survival probabilities for all-cause mortality. Full health cross-classification, self-rated health and health status. 89 observations were above graph maximum of age 83.

Results from Cox proportional hazards Model 1-3 are presented in Table 3. Compared to excellent self-rated health and favourable health status, all other levels of the full health cross-classification were associated with greater hazards, ranging from HR = 1.22 (95% CI 1.15-1.29, *p*_Bonf._ < 0.001) for good self-rated health and favourable health status to HR = 7.14 (95% CI 6.70-7.60, *p*_Bonf._ < 0.001) for poor self-rated health and unfavourable health status. The order of these levels was consistent across the three models, suggesting that individuals with concordant favourable self-rated health and health status had the lowest hazard and those with concordant unfavourable self-rated health and health status had the highest hazard. Participants with discordant self-rated health and health status had intermediate hazards. Compared to the results from Model 1, we observed attenuation in the hazard ratios from Model 2 and 3, although the overall pattern of results was consistent across these analyses (all Bonferroni-adjusted *p*-values < 0.001). Survival probabilities and cumulative hazards for Model 3 are presented in Figure 4.

**Table 3.** Cox proportional hazards model: all-cause mortality

|  |  | Model 1 |  |  | Model 2 |  |  | Model 3 |  |  |
| --- | --- | --- | --- | --- | --- | --- | --- | --- | --- | --- |
| Health cross-classification |  | HR | 95% CI |  | HR | 95% CI |  | HR | 95% CI |  |
| Excellent | Favourable | Ref | - | - | Ref | - | - | Ref | - | - |
| Good | Favourable | 1.22 | 1.15 | 1.29 | 1.20 | 1.14 | 1.27 | 1.16 | 1.10 | 1.23 |
| Excellent | Unfavourable | 1.42 | 1.30 | 1.55 | 1.38 | 1.26 | 1.51 | 1.37 | 1.25 | 1.50 |
| Fair | Favourable | 1.86 | 1.74 | 1.99 | 1.77 | 1.65 | 1.89 | 1.59 | 1.49 | 1.70 |
| Good | Unfavourable | 1.87 | 1.77 | 1.98 | 1.81 | 1.71 | 1.91 | 1.73 | 1.63 | 1.83 |
| Poor | Favourable | 2.79 | 2.42 | 3.22 | 2.62 | 2.27 | 3.02 | 2.17 | 1.88 | 2.51 |
| Fair | Unfavourable | 3.26 | 3.08 | 3.45 | 3.02 | 2.85 | 3.19 | 2.72 | 2.57 | 2.88 |
| Poor | Unfavourable | 7.14 | 6.70 | 7.60 | 6.34 | 5.94 | 6.76 | 5.24 | 4.90 | 5.61 |

Note: HR = hazard ratio; CI = confidence interval. Model 1 – unadjusted; Model 2 – adjusted for sociodemographic characteristics; Model 3 – additionally adjusted for lifestyle factors. All Bonferroni-adjusted $p$ -values < 0.001.

**Figure 4.**
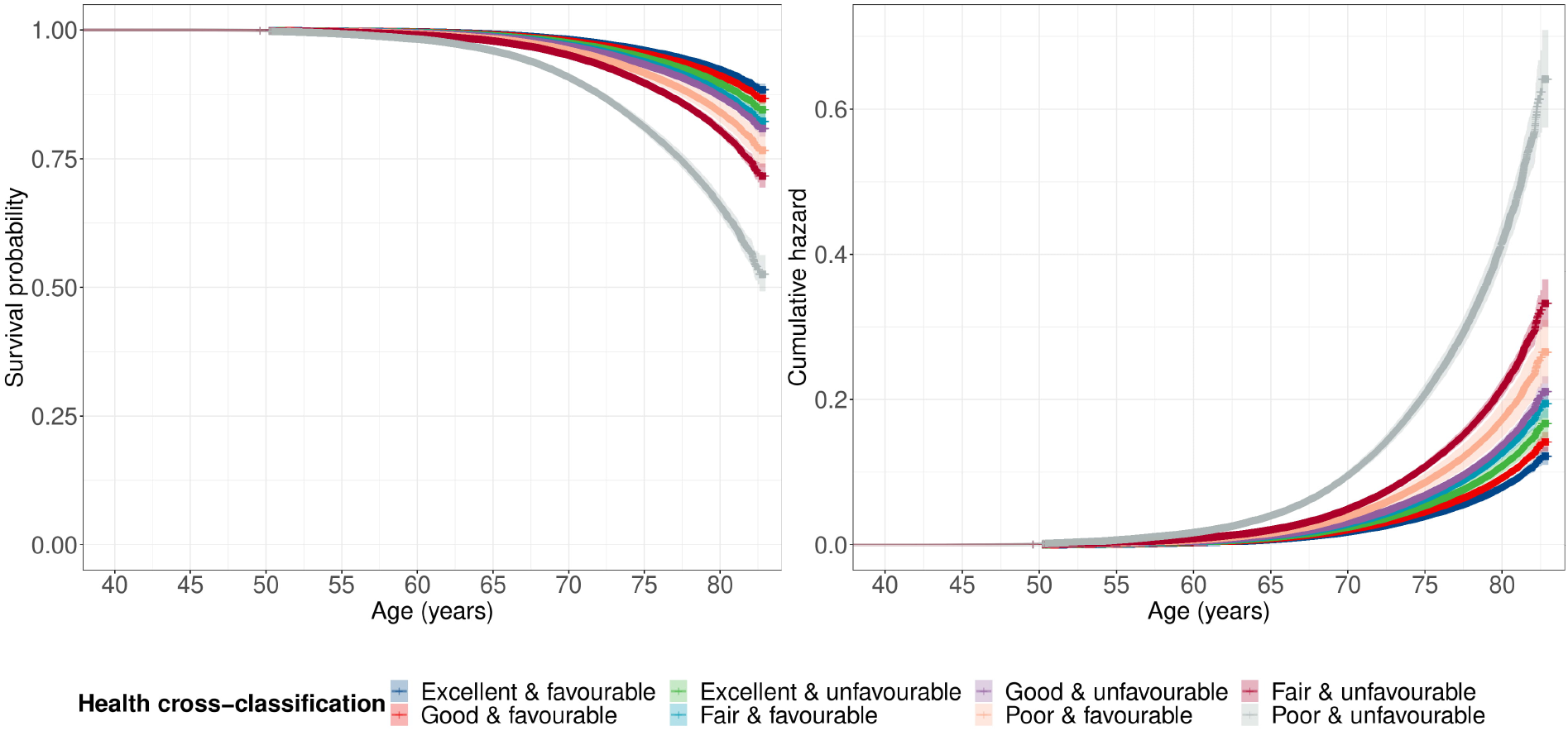
Cox proportional hazard survival probabilities and cumulative hazards for all-cause mortality. Estimates from Model 3 adjusted for sex, ethnicity, highest qualification, household income, smoking status, alcohol intake and physical activity. 89 observations were above graph maximum of age 83. Figures show continuous covariates fixed at their mean value and categorical covariates fixed at the largest group.

### Cause-specific mortality

Survival probabilities for specific causes of death are presented in **Supplement Figure 3**. Results from Cox proportional hazards Model 1-3 are presented in Table 4 and **Supplement Table 2**. Compared to participants with excellent self-rated health and favourable health status, all other levels of the health cross-classification were associated with higher hazards. For example, HRs for ischaemic heart disease from Model 3 ranged from HR = 1.14 (95% CI 0.97-1.35, *p*_BH_ = 0.12) for good self-rated health and favourable health status to HR = 5.17 (95% CI 4.17-6.26, *p*_Bonf._ < 0.001) for poor self-rated health and unfavourable health status. We generally observed attenuation in the hazard ratios from Model 2 and 3, compared to those from Model 1. For example, the highest hazard ratio for malignant neoplasm was HR = 8.10 (95% CI 5.51-11.9, *p*_Bonf._ < 0.001) for poor self-rated health and unfavourable health status in Model 1, HR = 6.96 (95% CI 4.71-10.31, *p*_Bonf._ < 0.001) in Model 2 and HR = 5.80 (95% CI 3.87-8.68, *p*_Bonf._ < 0.001) in Model 3. The overall pattern of results was similar to the results for all-cause mortality, although several comparisons were not statistically significant after multiple testing correction, typically for causes of death with lower numbers of observed deaths (influenza and pneumonia, dementia and Alzheimer’s disease, and chronic lower respiratory disease) and levels of the health cross-classification that were associated with longer survival times than good self-rated health and unfavourable health status.

**Table 4.**
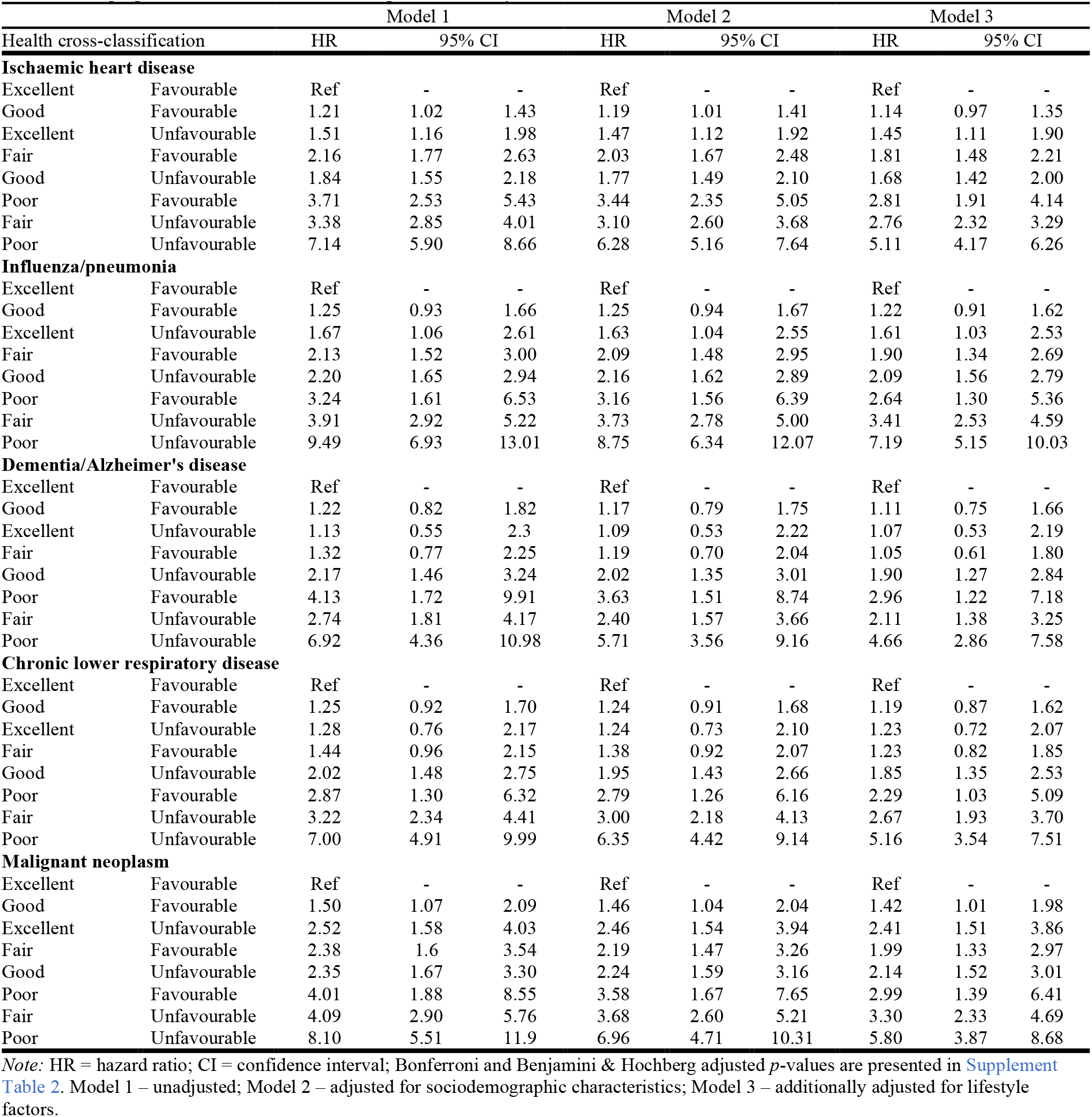
Cox proportional hazards model: cause-specific mortality

|  |  | Model 1 |  |  | Model 2 |  |  | Model 3 |  |  |
| --- | --- | --- | --- | --- | --- | --- | --- | --- | --- | --- |
| Health cross-classification |  | HR | 95% CI |  | HR | 95% CI |  | HR | 95% CI |  |
| **Ischaemic heart disease** |  |  |  |  |  |  |  |  |  |  |
| Excellent | Favourable | Ref | - | - | Ref | - | - | Ref | - | - |
| Good | Favourable | 1.21 | 1.02 | 1.43 | 1.19 | 1.01 | 1.41 | 1.14 | 0.97 | 1.35 |
| Excellent | Unfavourable | 1.51 | 1.16 | 1.98 | 1.47 | 1.12 | 1.92 | 1.45 | 1.11 | 1.90 |
| Fair | Favourable | 2.16 | 1.77 | 2.63 | 2.03 | 1.67 | 2.48 | 1.81 | 1.48 | 2.21 |
| Good | Unfavourable | 1.84 | 1.55 | 2.18 | 1.77 | 1.49 | 2.10 | 1.68 | 1.42 | 2.00 |
| Poor | Favourable | 3.71 | 2.53 | 5.43 | 3.44 | 2.35 | 5.05 | 2.81 | 1.91 | 4.14 |
| Fair | Unfavourable | 3.38 | 2.85 | 4.01 | 3.10 | 2.60 | 3.68 | 2.76 | 2.32 | 3.29 |
| Poor | Unfavourable | 7.14 | 5.90 | 8.66 | 6.28 | 5.16 | 7.64 | 5.11 | 4.17 | 6.26 |
| **Influenza/pneumonia** |  |  |  |  |  |  |  |  |  |  |
| Excellent | Favourable | Ref | - | - | Ref | - | - | Ref | - | - |
| Good | Favourable | 1.25 | 0.93 | 1.66 | 1.25 | 0.94 | 1.67 | 1.22 | 0.91 | 1.62 |
| Excellent | Unfavourable | 1.67 | 1.06 | 2.61 | 1.63 | 1.04 | 2.55 | 1.61 | 1.03 | 2.53 |
| Fair | Favourable | 2.13 | 1.52 | 3.00 | 2.09 | 1.48 | 2.95 | 1.90 | 1.34 | 2.69 |
| Good | Unfavourable | 2.20 | 1.65 | 2.94 | 2.16 | 1.62 | 2.89 | 2.09 | 1.56 | 2.79 |
| Poor | Favourable | 3.24 | 1.61 | 6.53 | 3.16 | 1.56 | 6.39 | 2.64 | 1.30 | 5.36 |
| Fair | Unfavourable | 3.91 | 2.92 | 5.22 | 3.73 | 2.78 | 5.00 | 3.41 | 2.53 | 4.59 |
| Poor | Unfavourable | 9.49 | 6.93 | 13.01 | 8.75 | 6.34 | 12.07 | 7.19 | 5.15 | 10.03 |
| **Dementia/Alzheimer's disease** |  |  |  |  |  |  |  |  |  |  |
| Excellent | Favourable | Ref | - | - | Ref | - | - | Ref | - | - |
| Good | Favourable | 1.22 | 0.82 | 1.82 | 1.17 | 0.79 | 1.75 | 1.11 | 0.75 | 1.66 |
| Excellent | Unfavourable | 1.13 | 0.55 | 2.3 | 1.09 | 0.53 | 2.22 | 1.07 | 0.53 | 2.19 |
| Fair | Favourable | 1.32 | 0.77 | 2.25 | 1.19 | 0.70 | 2.04 | 1.05 | 0.61 | 1.80 |
| Good | Unfavourable | 2.17 | 1.46 | 3.24 | 2.02 | 1.35 | 3.01 | 1.90 | 1.27 | 2.84 |
| Poor | Favourable | 4.13 | 1.72 | 9.91 | 3.63 | 1.51 | 8.74 | 2.96 | 1.22 | 7.18 |
| Fair | Unfavourable | 2.74 | 1.81 | 4.17 | 2.40 | 1.57 | 3.66 | 2.11 | 1.38 | 3.25 |
| Poor | Unfavourable | 6.92 | 4.36 | 10.98 | 5.71 | 3.56 | 9.16 | 4.66 | 2.86 | 7.58 |
| **Chronic lower respiratory disease** |  |  |  |  |  |  |  |  |  |  |
| Excellent | Favourable | Ref | - | - | Ref | - | - | Ref | - | - |
| Good | Favourable | 1.25 | 0.92 | 1.70 | 1.24 | 0.91 | 1.68 | 1.19 | 0.87 | 1.62 |
| Excellent | Unfavourable | 1.28 | 0.76 | 2.17 | 1.24 | 0.73 | 2.10 | 1.23 | 0.72 | 2.07 |
| Fair | Favourable | 1.44 | 0.96 | 2.15 | 1.38 | 0.92 | 2.07 | 1.23 | 0.82 | 1.85 |
| Good | Unfavourable | 2.02 | 1.48 | 2.75 | 1.95 | 1.43 | 2.66 | 1.85 | 1.35 | 2.53 |
| Poor | Favourable | 2.87 | 1.30 | 6.32 | 2.79 | 1.26 | 6.16 | 2.29 | 1.03 | 5.09 |
| Fair | Unfavourable | 3.22 | 2.34 | 4.41 | 3.00 | 2.18 | 4.13 | 2.67 | 1.93 | 3.70 |
| Poor | Unfavourable | 7.00 | 4.91 | 9.99 | 6.35 | 4.42 | 9.14 | 5.16 | 3.54 | 7.51 |
| **Malignant neoplasm** |  |  |  |  |  |  |  |  |  |  |
| Excellent | Favourable | Ref | - | - | Ref | - | - | Ref | - | - |
| Good | Favourable | 1.50 | 1.07 | 2.09 | 1.46 | 1.04 | 2.04 | 1.42 | 1.01 | 1.98 |
| Excellent | Unfavourable | 2.52 | 1.58 | 4.03 | 2.46 | 1.54 | 3.94 | 2.41 | 1.51 | 3.86 |
| Fair | Favourable | 2.38 | 1.6 | 3.54 | 2.19 | 1.47 | 3.26 | 1.99 | 1.33 | 2.97 |
| Good | Unfavourable | 2.35 | 1.67 | 3.30 | 2.24 | 1.59 | 3.16 | 2.14 | 1.52 | 3.01 |
| Poor | Favourable | 4.01 | 1.88 | 8.55 | 3.58 | 1.67 | 7.65 | 2.99 | 1.39 | 6.41 |
| Fair | Unfavourable | 4.09 | 2.90 | 5.76 | 3.68 | 2.60 | 5.21 | 3.30 | 2.33 | 4.69 |
| Poor | Unfavourable | 8.10 | 5.51 | 11.9 | 6.96 | 4.71 | 10.31 | 5.80 | 3.87 | 8.68 |

Note: HR = hazard ratio; CI = confidence interval; Bonferroni and Benjamini & Hochberg adjusted *p*-values are presented in Supplement Table 2. Model 1 – unadjusted; Model 2 – adjusted for sociodemographic characteristics; Model 3 – additionally adjusted for lifestyle factors.

### Sensitivity analysis

The average number of illnesses was 1.07 (SD = 1.22) in individuals with good/excellent self-rated health and favourable health status, 2.78 (SD = 1.67) in individuals with good/excellent self-rated health and unfavourable health status, 1.86 (SD = 1.59) in individuals with poor/fair self-rated health and favourable health status and 4.00 (SD = 2.30) in individuals with poor/fair self-rated health and unfavourable health status.

Kaplan-Meier survival probabilities from our sensitivity analysis are presented in Figure 5, suggesting that different levels of the simplified health cross-classification were associated with varying survival times (log-rank test *p* < 0.001). We found that participants with favourable health status and good or excellent self-rated health had the longest survival times, while participants with unfavourable health status and poor or fair self-rated health had the shortest survival times. Participants with discordant health status and self-rated health had intermediate survival times. Individuals with good or excellent self-rated health but unfavourable health status, i.e., for whom their objective assessment of health was worse, had only slightly shorter survival times than participants with favourable health status but fair or poor self-rated health.

**Figure 5.**
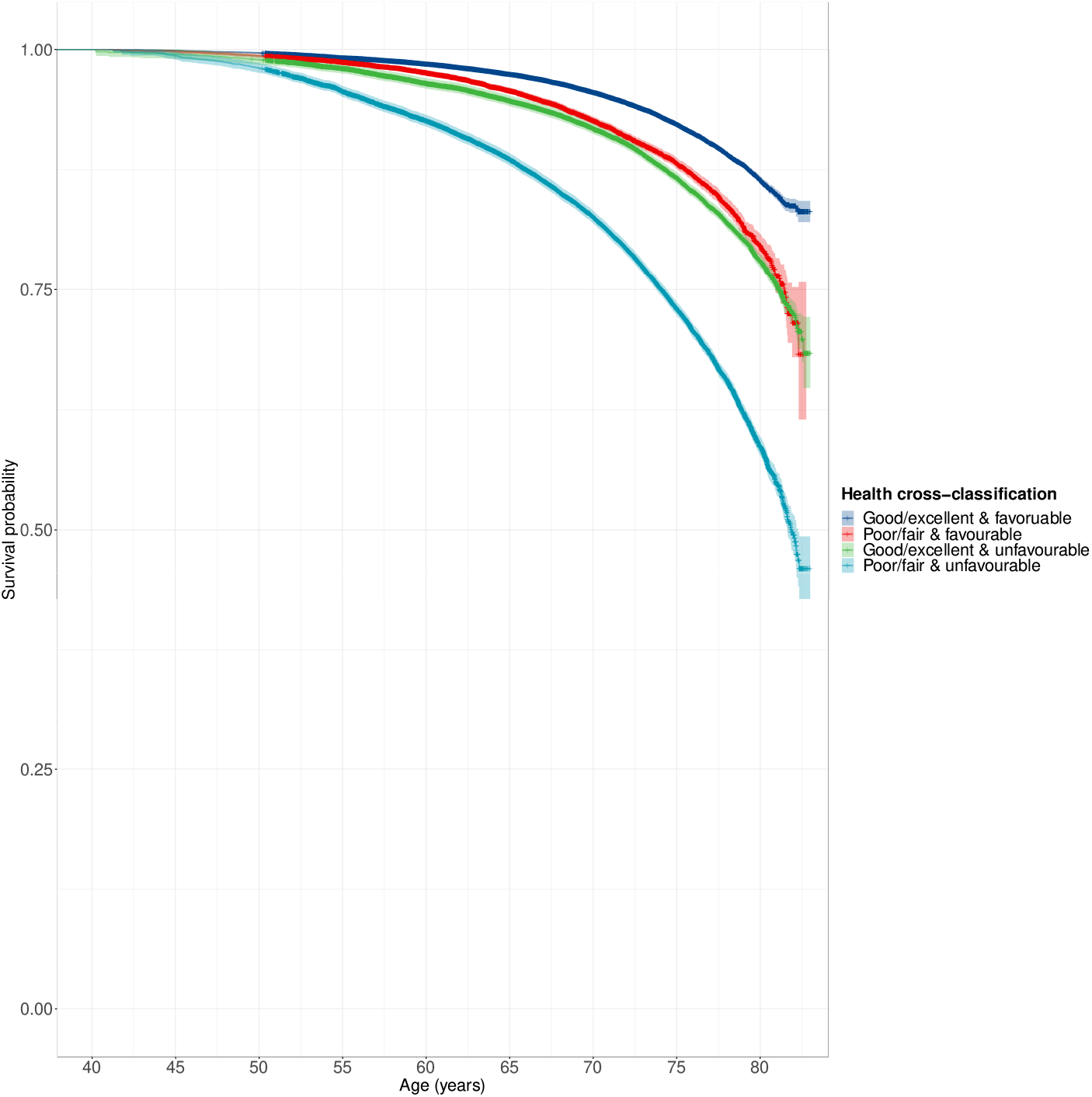
Kaplan-Meier survival probabilities for all-cause mortality. Simplified health cross-classification. 89 observations were above graph maximum of age 83.

Results from Cox proportional hazards Model 1-3 are presented in Table 5. Compared to favourable health status and good or excellent self-rated health, all other levels of the simplified health cross-classification were associated with increased hazards. Across Model 1 and 2, individuals with better self-rated health than health status had lower hazards than individuals with worse self-rated health than health status, compared to the reference group, although in Model 3 those with worse self-rated health than health status had lower hazards, similar to the Kaplan-Meier estimates. Individuals with unfavourable health status and self-rated health had the highest hazards across all models.

**Table 5.** Cox proportional hazards model: all-cause mortality

| Table 5. Cox proportional hazards model: all-cause mortality |  |  |  |  |  |  |  |  |  |  |
| --- | --- | --- | --- | --- | --- | --- | --- | --- | --- | --- |
|  |  | Model 1 |  |  | Model 2 |  |  | Model 3 |  |  |
| Term |  | HR | 95% CI |  | HR | 95% CI |  | HR | 95% CI |  |
| Good/excellent | Favourable | Ref | - | - | Ref | - | - | Ref | - | - |
| Good/excellent | Unfavourable | 1.55 | 1.50 | 1.61 | 1.51 | 1.46 | 1.56 | 1.49 | 1.44 | 1.54 |
| Poor/fair | Favourable | 1.66 | 1.58 | 1.75 | 1.58 | 1.50 | 1.66 | 1.43 | 1.36 | 1.50 |
| Poor/fair | Unfavourable | 3.43 | 3.32 | 3.54 | 3.14 | 3.04 | 3.25 | 2.81 | 2.72 | 2.91 |

Note: HR = hazard ratio; CI = confidence interval. Model 1 – unadjusted; Model 2 – adjusted for sociodemographic characteristics; Model 3 – additionally adjusted for lifestyle factors. All Bonferroni-adjusted $p$ -values < 0.001.

## Discussion

Using data from more than 370,000 UK Biobank participants with a median follow-up period of almost 12 years, we found that individuals with favourable health status or good to excellent self-rated health at baseline had a substantially lower risk of mortality than individuals with unfavourable health status or poor to fair self-rated health, respectively. Examining the cross-classification between health status and self-rated health provided additional granularity to differentiate survival times, confirming that self-rated health captures additional health-related information and highlighting the potential value of combining these measures for risk stratification.

### Principal findings

As hypothesised, individuals with favourable health status and good to excellent self-rated health had the longest survival times, while individuals with unfavourable health status and poor to fair self-rated health had the shortest survival times. Individuals with discordant health status and self-rated health had intermediate survival times. For example, individuals with favourable health status based on their medical history but poor self-rated health had substantially shorter survival times than individuals with favourable health status and good to excellent self-rated health.

We observed similar results for leading causes of death in the UK^6^. However, there were some inconsistencies for causes of death with fewer observed deaths during the follow-up period. For dementia and Alzheimer’s disease, we did not find evidence of differences between the four health cross-classification levels with the longest survival times: (i) good self-rated health and favourable health status, (ii) excellent self-rated health and unfavourable health status and (iii) fair self-rated health and favourable health status, compared to (iv) excellent self-rated health and favourable health status.

In the sensitivity analysis in which we merged good and excellent self-rated health and poor and fair self-rated health prior to analysis, we observed that individuals with discordant health status and self-rated health had intermediate survival times, showing that both measures have a key contribution to predicting all-cause mortality.

### Findings in context

A previous analysis of 5-year mortality in the UK Biobank identified self-rated health as the strongest predictor of all-cause mortality in men, out of 655 variables, and showed that self-rated health was consistently associated with cause-specific mortality^12^. Our findings support most previous research that found a graded association between self-rated health levels and mortality^1^. Findings from a Brazilian cohort study suggested that the 10-year mortality risk was similar for participants who reported fair or good to excellent self-rated health, compared to poor self-rated health^4^. However, the sample size was limited and the difference in result might be due to linguistic factors; ‘fair’ self-rated health might reflect average or normal health in Spanish or Portugese^17^, while it might reflect less favourable health in English. Although several previous studies have examined mortality outcomes associated with objective and subjective assessments of health^3,4^, no studies have, to our knowledge, examined the cross-classification between health status and self-rated health for potential risk stratification.

### Strengths and limitations

Strengths of this study include its large sample size (>370,00 participants) and a median follow-up of almost 12 years. Risk indices are often developed in high-risk populations (e.g., in older individuals), focus on a single health outcome and studies are often limited by small sample sizes. The health cross-classification examined in this study might be applicable for risk stratification for a wide range of health outcomes.

Our research inevitably has limitations. There might be some misclassification in the reporting of medical illnesses that were used to determine health status. However, participants were asked to report illnesses that had been diagnosed by a doctor and these diagnoses were confirmed during a nurse-led interview. Nevertheless, there is the possibility of recall bias in the reporting of long-term and past conditions. Regarding self-rated health, there could be differences by native language in evaluating fair self-rated health as positive or negative. However, most participants were born in the UK and such differences, if present, would likely be minimal. Mortality data from death certificates might have some misclassification in causes of death, especially between similar diseases, and we observed fewer deaths during follow-up for several specific causes of death. However, all-cause mortality is a more robust endpoint than cause-specific mortality and we found similar results across most outcomes.

### Generalisability

Compared with non-responders, UK Biobank participants were older, more likely to be female and more likely of higher socioeconomic status. They were also less likely to engage in unhealthy lifestyle behaviours, reported fewer medical illnesses compared with data from a nationally representative survey and all-cause mortality was 46.2% lower for 70-74-year-olds^18^. Although these findings show that the UK Biobank is not fully representative of the UK population, a recent empirical investigation comparing the UK Biobank with data from 18 prospective cohort studies with conventional response rates showed that the direction of risk factor associations were similar, although with differences in magnitude^19^. It is also worth noting that most participants (79.6%) in our sample reported good to excellent self-rated health, which is comparable to estimates from the Office for National Statistics suggesting that >72% of people in England and Wales rated their health as good or very good^20^.

### Implications

Risk stratification is a key public health priority that is central to clinical decision making and resource allocation. Self-rated health and health status can be obtained through verbal interview, self-report or from medical records. The cross-classification between health status and self-rated health represents a straightforward metric for initial risk stratification, with applications to population health, clinical decision making and resource allocation. Our findings also highlight that self-rated health captures additional health-related information and should therefore be more widely assessed across settings.

## Data Availability

The data used are available to all bona fide researchers for health-related research that is in the public interest, subject to an application process and approval criteria. Study materials are publicly available online at http://www.ukbiobank.ac.uk.

## Authorship contributions

CML acquired the studentship funding, interpreted the findings and critically reviewed the manuscript. JM conceived the idea of the study, acquired the data, carried out the statistical analysis, interpreted the findings, wrote the manuscript and revised the manuscript for final submission. Both authors read and approved the final manuscript. JM had full access to all data used in this study and takes responsibility for the integrity of the data and the accuracy of the data analysis.

## Acknowledgments

This research has been conducted using data from UK Biobank, a major biomedical database. This project made use of time on Rosalind HPC, funded by Guy’s & St Thomas’ Hospital NHS Trust Biomedical Research Centre (GSTT-BRC), South London & Maudsley NHS Trust Biomedical Research Centre (SLAM-BRC), and Faculty of Natural Mathematics & Science (NMS) at King’s College London.

## Ethics

Ethical approval for the UK Biobank study has been granted by the National Information Governance Board for Health and Social Care and the NHS North West Multicentre Research Ethics Committee (11/NW/0382). No project-specific ethical approval is needed. Data access permission has been granted under UK Biobank application 45514.

## Supplementary material

Supplementary information is available online.

