## Supplement for "Cross-classification between self-rated health and health status: longitudinal analyses of all-cause mortality and leading causes of death in the UK"

### Supplement 1: Hypotheses

#### Full health cross-classification

Full health cross-classification

| | Self-rated health | | | |
|---|---|---|---|---|
| Health status | Poor | Fair | Good | Excellent |
| Favourable | ? | ? | ↑ | ↑↑ |
| Unfavourable | ↓↓ | ↓ | ? | ? |

#### Reduced health cross-classification

Reduced health cross-classification

| | Self-rated health | |
|---|---|---|
| Health status | Poor/Fair | Good/Excellent |
| Favourable | ? | ↑ |
| Unfavourable | ↓ | ? |

### Supplement 2: Additional results

#### Supplement table 1. Sample characteristics

**Supplement table 1**. Sample characteristics

| | Full sample (N=502521) | Analytical sample (N=373761) |
|---|---|---|
| **Age** | | |
| Mean (SD) | 56.53 (8.10) | 56.02 (8.08) |
| **Sex** | | |
| Female | 273394 (54.4%) | 193541 (51.8%) |
| Male | 229127 (45.6%) | 180220 (48.2%) |
| **Ethnicity** | | |
| White | 472711 (94.1%) | 357509 (95.7%) |
| Mixed-race | 2958 (0.6%) | 2117 (0.6%) |
| Black | 8061 (1.6%) | 4877 (1.3%) |
| Asian | 9882 (2.0%) | 5551 (1.5%) |
| Chinese | 1574 (0.3%) | 1023 (0.3%) |
| Other | 4558 (0.9%) | 2684 (0.7%) |
| Prefer not to answer | 1662 (0.3%) | |
| Do not know | 217 (0.0%) | |
| Missing | 898 (0.2%) | |
| **Highest qualification**[1] | | |
| None | 85274 (17.0%) | 49621 (13.3%) |
| O levels/GCSEs/CSEs | 132086 (26.3%) | 99810 (26.7%) |
| A levels/NVQ/HND/HNC[1] | 113859 (22.7%) | 88033 (23.6%) |
| Degree | 161168 (32.1%) | 136297 (36.5%) |
| Prefer not to answer | 5493 (1.1%) | |
| Missing | 4641 (0.9%) | |
| **Household income**[2] | | |
| Very low | 97205 (19.3%) | 78241 (20.9%) |
| Low | 108177 (21.5%) | 94386 (25.3%) |
| Medium | 110774 (22.0%) | 99882 (26.7%) |
| High | 86269 (17.2%) | 79742 (21.3%) |
| Very high | 22930 (4.6%) | 21510 (5.8%) |
| Prefer not to answer | 49848 (9.9%) | |
| Do not know | 21305 (4.2%) | |
| Missing | 6013 (1.2%) | |
| **Walking** (days/week)[3] | | |
| Mean (SD) | 5.39 (1.93) | 5.36 (1.95) |
| Prefer not to answer | 979 (0.19%) | |
| Do not know | 6687 (1.33%) | |
| Unable to walk | 1929 (0.38%) | |
| Missing | 874 (0.17%) | |
| **Moderate activity** (days/week)[3] | | |
| Mean (SD) | 3.63 (2.33) | 3.57 (2.33) |
| Prefer not to answer | 2273 (0.45%) | |
| Do not know | 24120 (4.8%) | |
| Missing | 878 (0.17%) | |
| **Vigorous activity** (days/week)[3] | | |
| Mean (SD) | 1.84 (1.96) | 1.86 (1.94) |
| Prefer not to answer | 4116 (0.82%) | |
| Do not know | 22582 (4.49%) | |
| Missing | 878 (0.17%) | |
| **Smoking status** | | |
| Never | 273528 (54.4%) | 204457 (54.7%) |
| Former | 173064 (34.4%) | 131186 (35.1%) |
| Current | 52979 (10.5%) | 38118 (10.2%) |
| Prefer not to answer | 2059 (0.4%) | |
| Missing | 891 (0.2%) | |
| **Alcohol intake frequency** | | |
| Never | 40645 (8.1%) | 25358 (6.8%) |
| Special occasions | 58011 (11.5%) | 38719 (10.4%) |
| 1-3/month | 55856 (11.1%) | 41310 (11.1%) |
| 1-2/week | 129294 (25.7%) | 96316 (25.8%) |
| 3-4/week | 115443 (23.0%) | 91046 (24.4%) |
| Daily/almost daily | 101770 (20.3%) | 81012 (21.7%) |
| Prefer not to answer | 605 (0.1%) | |
| Missing | 897 (0.2%) | |

*Note:* SD = standard deviation; GCSEs = general certificate of secondary education; CSE = certificate of secondary education; NVQ = national vocational qualification; HND = higher national diploma; HNC = higher national certificate. [1]also includes 'other professional qualifications'. [2]Annual household income groups: very low (<£18 000), low (£18 000–30 999), middle (£31 000–51 999), high (£52 000–100 000) and very high (>£100 000). [3]number of days per week engaging in these activities for 10+ minutes continuously.

Supplement figure 1. Non-cancer illnesses

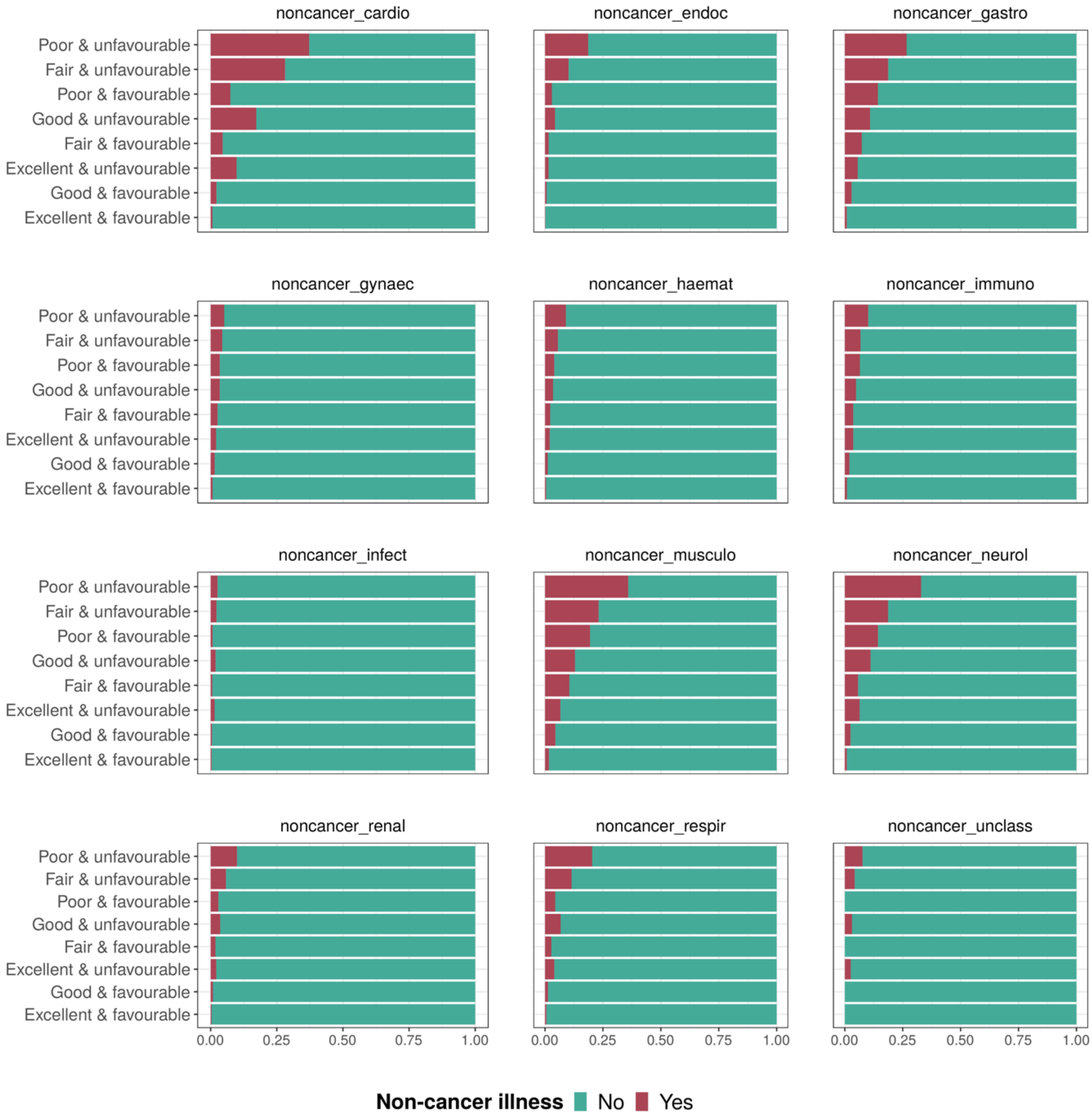

**Supplement figure 1.** Non-cancer illnesses by health cross-classification. Proportions represent individuals with at least one non-cancer illness within each broader illness group.

Supplement figure 2. Cancer illnesses

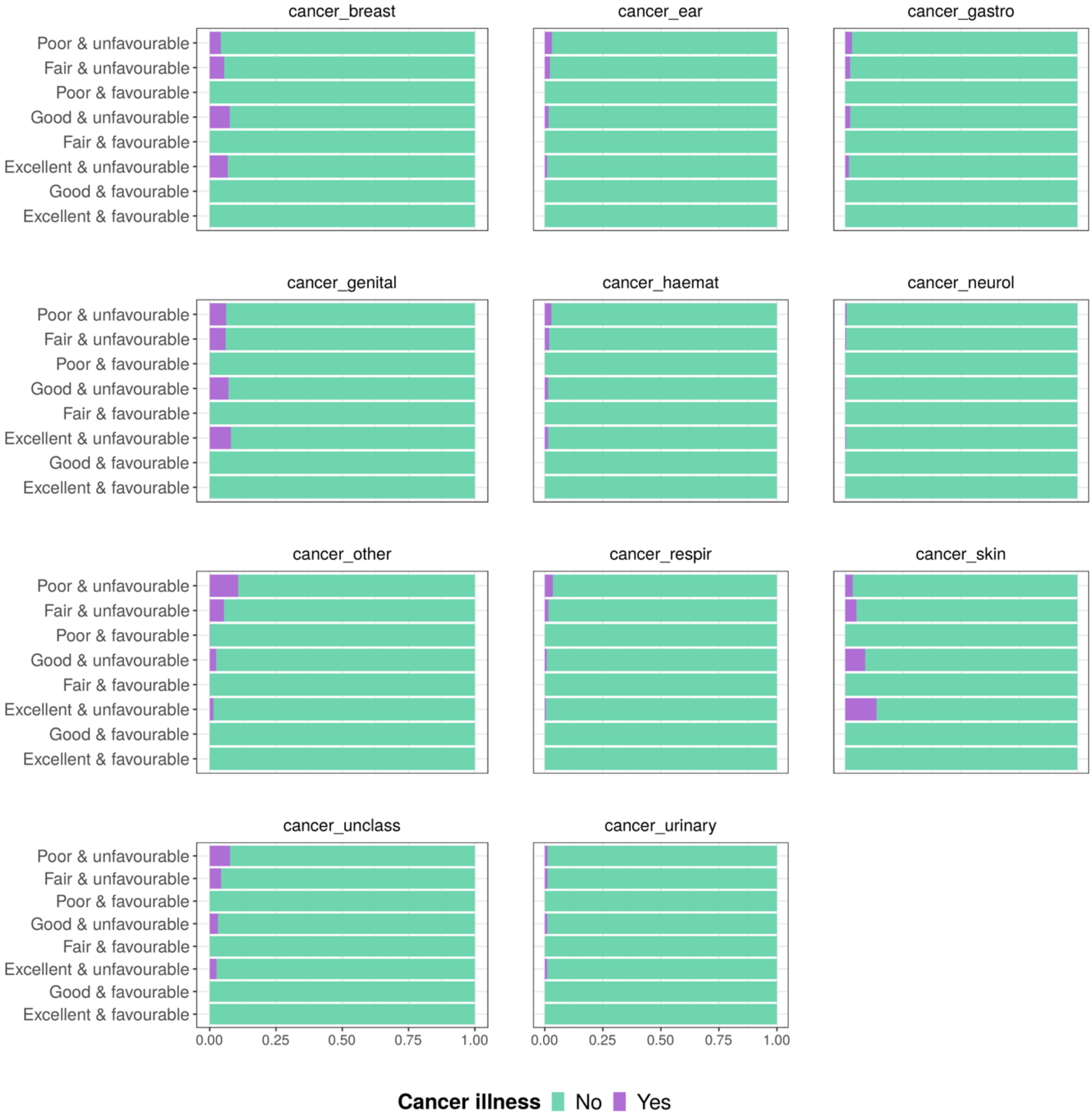

**Supplement figure 2.** Cancer illnesses by health cross-classification. Proportions represent individuals with at least one cancer illness within each broader illness group.

Supplement figure 3. Survival curves cause-specific mortality

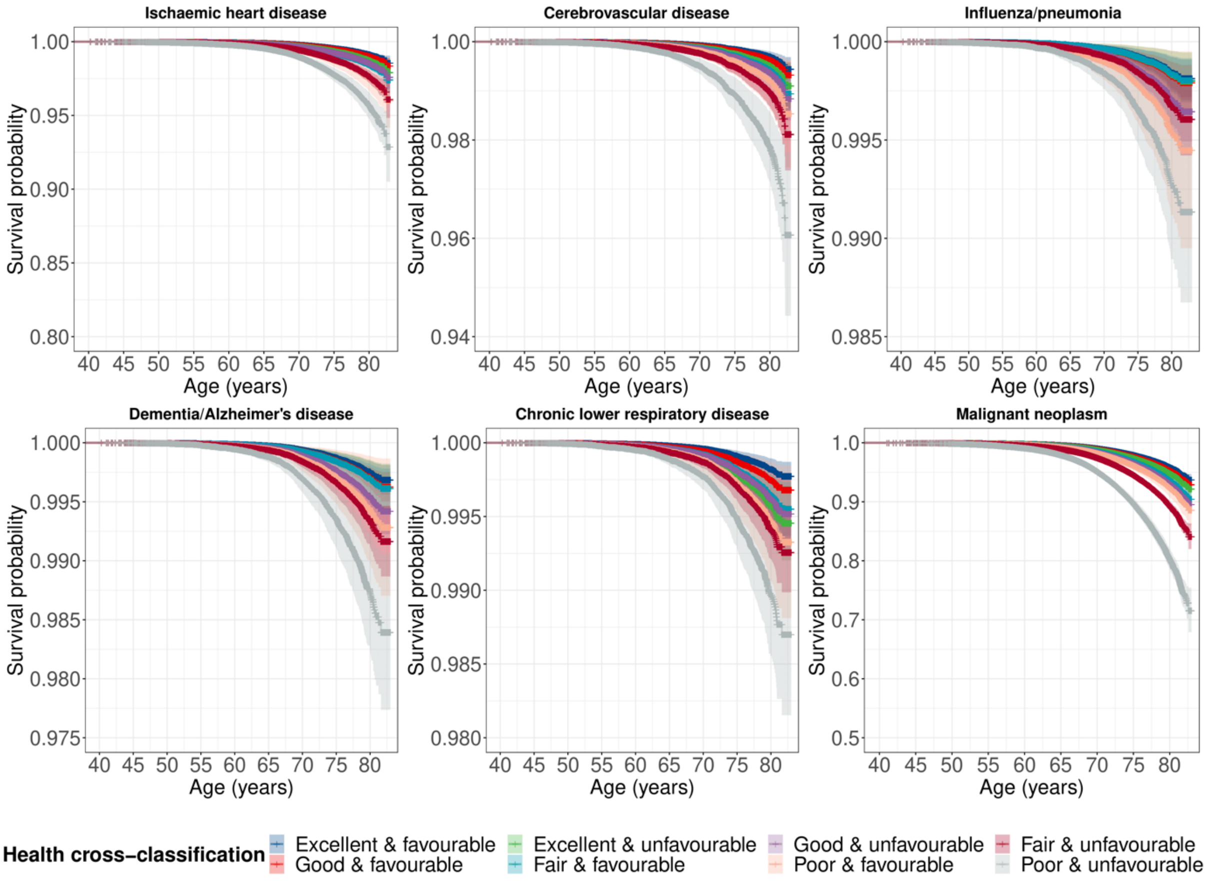

**Supplement figure 3.** Cox proportional hazard survival curves for cause-specific mortality.

Supplement table 2. Hazard ratios cause specific mortality

Results presented below correspond to Table 4 presented in the main body of the text but also include $p$-values. Results for which Bonferroni and Benjamini & Hochberg adjusted $p$-values differ are highlighted.

**Supplement table 2.** Cox proportional hazards model: cause-specific mortality

| Term | | Model 1 | | | | | Model 2 | | | | | Model 3 | | | | |
|---|---|---|---|---|---|---|---|---|---|---|---|---|---|---|---|---|
| | | HR | 95% CI | | $p_{\text{Bonf.}}$ | $p_{\text{BH}}$ | HR | 95% CI | | $p_{\text{Bonf.}}$ | $p_{\text{BH}}$ | HR | 95% CI | | $p_{\text{Bonf.}}$ | $p_{\text{BH}}$ |
| **Ischaemic heart disease** | | | | | | | | | | | | | | | | |
| Excellent | Favourable | Ref | - | - | - | - | Ref | - | - | - | - | Ref | - | - | - | - |
| Good | Favourable | 1.21 | 1.02 | 1.43 | 0.5566 | 0.0293 | 1.19 | 1.01 | 1.41 | 0.8886 | 0.0444 | 1.14 | 0.97 | 1.35 | >0.999 | 0.1219 |
| Excellent | Unfavourable | 1.51 | 1.16 | 1.98 | 0.0541 | 0.0034 | 1.47 | 1.12 | 1.92 | 0.1054 | 0.0062 | 1.45 | 1.11 | 1.90 | 0.135 | 0.0075 |
| Fair | Favourable | 2.16 | 1.77 | 2.63 | <0.001 | <0.001 | 2.03 | 1.67 | 2.48 | <0.001 | <0.001 | 1.81 | 1.48 | 2.21 | <0.001 | <0.001 |
| Good | Unfavourable | 1.84 | 1.55 | 2.18 | <0.001 | <0.001 | 1.77 | 1.49 | 2.10 | <0.001 | <0.001 | 1.68 | 1.42 | 2.00 | <0.001 | <0.001 |
| Poor | Favourable | 3.71 | 2.53 | 5.43 | <0.001 | <0.001 | 3.44 | 2.35 | 5.05 | <0.001 | <0.001 | 2.81 | 1.91 | 4.14 | <0.001 | <0.001 |
| Fair | Unfavourable | 3.38 | 2.85 | 4.01 | <0.001 | <0.001 | 3.10 | 2.60 | 3.68 | <0.001 | <0.001 | 2.76 | 2.32 | 3.29 | <0.001 | <0.001 |
| Poor | Unfavourable | 7.14 | 5.90 | 8.66 | <0.001 | <0.001 | 6.28 | 5.16 | 7.64 | <0.001 | <0.001 | 5.11 | 4.17 | 6.26 | <0.001 | <0.001 |
| **Influenza/pneumonia** | | | | | | | | | | | | | | | | |
| Excellent | Favourable | Ref | - | - | - | - | Ref | - | - | - | - | Ref | - | - | - | - |
| Good | Favourable | 1.25 | 0.93 | 1.66 | >0.999 | 0.1398 | 1.25 | 0.94 | 1.67 | >0.999 | 0.1398 | 1.22 | 0.91 | 1.62 | >0.999 | 0.1862 |
| Excellent | Unfavourable | 1.67 | 1.06 | 2.61 | 0.5500 | 0.0344 | 1.63 | 1.04 | 2.55 | 0.7101 | 0.0418 | 1.61 | 1.03 | 2.53 | 0.7822 | 0.0435 |
| Fair | Favourable | 2.13 | 1.52 | 3.00 | 0.0003 | <0.001 | 2.09 | 1.48 | 2.95 | 0.0006 | 0.0001 | 1.90 | 1.34 | 2.69 | 0.0061 | 0.0005 |
| Good | Unfavourable | 2.20 | 1.65 | 2.94 | <0.001 | <0.001 | 2.16 | 1.62 | 2.89 | <0.001 | <0.001 | 2.09 | 1.56 | 2.79 | <0.001 | <0.001 |
| Poor | Favourable | 3.24 | 1.61 | 6.53 | 0.0215 | 0.0017 | 3.16 | 1.56 | 6.39 | 0.0285 | 0.0020 | 2.64 | 1.30 | 5.36 | 0.1501 | 0.0100 |
| Fair | Unfavourable | 3.91 | 2.92 | 5.22 | <0.001 | <0.001 | 3.73 | 2.78 | 5.00 | <0.001 | <0.001 | 3.41 | 2.53 | 4.59 | <0.001 | <0.001 |
| Poor | Unfavourable | 9.49 | 6.93 | 13.01 | <0.001 | <0.001 | 8.75 | 6.34 | 12.07 | <0.001 | <0.001 | 7.19 | 5.15 | 10.03 | <0.001 | <0.001 |
| **Dementia/Alzheimer's disease** | | | | | | | | | | | | | | | | |
| Excellent | Favourable | Ref | - | - | - | - | Ref | - | - | - | - | Ref | - | - | - | - |
| SuGood | Favourable | 1.22 | 0.82 | 1.82 | >0.999 | 0.4835 | 1.17 | 0.79 | 1.75 | >0.999 | 0.6041 | 1.11 | 0.75 | 1.66 | >0.999 | 0.7403 |
| Excellent | Unfavourable | 1.13 | 0.55 | 2.3 | >0.999 | 0.8567 | 1.09 | 0.53 | 2.22 | >0.999 | 0.8567 | 1.07 | 0.53 | 2.19 | >0.999 | 0.8567 |
| Fair | Favourable | 1.32 | 0.77 | 2.25 | >0.999 | 0.4835 | 1.19 | 0.70 | 2.04 | >0.999 | 0.6835 | 1.05 | 0.61 | 1.80 | >0.999 | 0.8567 |
| Good | Unfavourable | 2.17 | 1.46 | 3.24 | 0.0029 | 0.0005 | 2.02 | 1.35 | 3.01 | 0.0125 | 0.0017 | 1.90 | 1.27 | 2.84 | 0.0367 | 0.0037 |
| Poor | Favourable | 4.13 | 1.72 | 9.91 | 0.0309 | 0.0034 | 3.63 | 1.51 | 8.74 | 0.0851 | 0.0077 | 2.96 | 1.22 | 7.18 | 0.3391 | 0.0283 |
| Fair | Unfavourable | 2.74 | 1.81 | 4.17 | <0.001 | <0.001 | 2.40 | 1.57 | 3.66 | 0.0011 | 0.0002 | 2.11 | 1.38 | 3.25 | 0.0133 | 0.0017 |
| Poor | Unfavourable | 6.92 | 4.36 | 10.98 | <0.001 | <0.001 | 5.71 | 3.56 | 9.16 | <0.001 | <0.001 | 4.66 | 2.86 | 7.58 | <0.001 | <0.001 |
| **Chronic lower respiratory disease** | | | | | | | | | | | | | | | | |
| Excellent | Favourable | Ref | - | - | - | - | Ref | - | - | - | - | Ref | - | - | - | - |
| Good | Favourable | 1.25 | 0.92 | 1.70 | >0.999 | 0.2193 | 1.24 | 0.91 | 1.68 | >0.999 | 0.2297 | 1.19 | 0.87 | 1.62 | >0.999 | 0.3373 |
| Excellent | Unfavourable | 1.28 | 0.76 | 2.17 | >0.999 | 0.3921 | 1.24 | 0.73 | 2.10 | >0.999 | 0.4400 | 1.23 | 0.72 | 2.07 | >0.999 | 0.4473 |
| Fair | Favourable | 1.44 | 0.96 | 2.15 | >0.999 | 0.1211 | 1.38 | 0.92 | 2.07 | >0.999 | 0.1787 | 1.23 | 0.82 | 1.85 | >0.999 | 0.3635 |
| Good | Unfavourable | 2.02 | 1.48 | 2.75 | 0.0002 | <0.001 | 1.95 | 1.43 | 2.66 | 0.0006 | 0.0001 | 1.85 | 1.35 | 2.53 | 0.0026 | 0.0003 |
| Poor | Favourable | 2.87 | 1.30 | 6.32 | 0.1856 | 0.0186 | 2.79 | 1.26 | 6.16 | 0.2350 | 0.0214 | 2.29 | 1.03 | 5.09 | 0.8615 | 0.0718 |
| Fair | Unfavourable | 3.22 | 2.34 | 4.41 | <0.001 | <0.001 | 3.00 | 2.18 | 4.13 | <0.001 | <0.001 | 2.67 | 1.93 | 3.70 | <0.001 | <0.001 |
| Poor | Unfavourable | 7.00 | 4.91 | 9.99 | <0.001 | <0.001 | 6.35 | 4.42 | 9.14 | <0.001 | <0.001 | 5.16 | 3.54 | 7.51 | <0.001 | <0.001 |
| **Malignant neoplasm** | | | | | | | | | | | | | | | | |
| Excellent | Favourable | Ref | - | - | - | - | Ref | - | - | - | - | Ref | - | - | - | - |
| Good | Favourable | 1.50 | 1.07 | 2.09 | 0.3816 | 0.0201 | 1.46 | 1.04 | 2.04 | 0.5837 | 0.0292 | 1.42 | 1.01 | 1.98 | 0.9040 | 0.0430 |
| Excellent | Unfavourable | 2.52 | 1.58 | 4.03 | 0.0023 | 0.0002 | 2.46 | 1.54 | 3.94 | 0.0034 | 0.0003 | 2.41 | 1.51 | 3.86 | 0.0049 | 0.0003 |
| Fair | Favourable | 2.38 | 1.6 | 3.54 | 0.0004 | <0.001 | 2.19 | 1.47 | 3.26 | 0.0025 | 0.0002 | 1.99 | 1.33 | 2.97 | 0.0168 | 0.0011 |
| Good | Unfavourable | 2.35 | 1.67 | 3.30 | <0.001 | <0.001 | 2.24 | 1.59 | 3.16 | 0.0001 | <0.001 | 2.14 | 1.52 | 3.01 | 0.0003 | <0.001 |
| Poor | Favourable | 4.01 | 1.88 | 8.55 | 0.0067 | 0.0004 | 3.58 | 1.67 | 7.65 | 0.0210 | 0.0012 | 2.99 | 1.39 | 6.41 | 0.1044 | 0.0058 |
| Fair | Unfavourable | 4.09 | 2.90 | 5.76 | <0.001 | <0.001 | 3.68 | 2.60 | 5.21 | <0.001 | <0.001 | 3.30 | 2.33 | 4.69 | <0.001 | <0.001 |
| Poor | Unfavourable | 8.10 | 5.51 | 11.9 | <0.001 | <0.001 | 6.96 | 4.71 | 10.31 | <0.001 | <0.001 | 5.80 | 3.87 | 8.68 | <0.001 | <0.001 |

*Note:* HR = hazard ratio; CI = confidence interval; Bonf. = Bonferroni; BH = Benjamini & Hochberg. Model 1 – unadjusted; Model 2 – adjusted for sociodemographic characteristics; Model 3 – additionally adjusted for lifestyle factors.
